# Placentas from healthy women living in a cutaneous leishmaniasis endemic region in Mexico harbor *Leishmania mexicana* parasites: Potential implications for pregnancy and neonatal outcomes

**DOI:** 10.64898/2026.05.11.26352970

**Authors:** Rodrigo X. Armijos, Brandon A. Berger, Alejandra González Ayala, Miriam A. Delgado-Hernandez, José Luis Acosta-Patiño, Esmelin Trinidad-Vázquez, Liliana Aline Fernández-Urrutia, Elizabeth Baz-Rojas, Javier Mancilla-Galindo, Candelaria Frías Selván, Juanita Ortíz-Avalos, Eva Cristina Avalos-Ortiz, Frida Häberle, Miriam Torres-Vasquez, M. Margaret Weigel, Allison H. Bartlett, Yuriria Paredes, Miroslava Avila-García, Magdalena Aguirre García, Norma Galindo-Sevilla

## Abstract

**Background:** Women living in leishmaniasis-endemic zones are regularly exposed to the sandfly vector in their environments. While case series and laboratory evidence consistently suggest transplacental transmission of *Leishmania* parasites with deleterious maternal-fetal effects, this issue has received insufficient attention, particularly in areas where the predominant *Leishmania* species is mainly associated with cutaneous disease.

**Methodology/Principal Findings:** We conducted an exploratory cross-sectional study in a highly endemic zone of Tabasco, Mexico, enrolling 53 women with singleton term deliveries between April 2018 and April 2020. Placental PCR was positive in 18 (34%) participants. Buccal swabs were positive in 11 (21.2%) of 52 newborns.

Immunofluorescence confirmed intracellular amastigotes within macrophages near the vascular endothelium of PCR-positive placentas, with no surrounding inflammatory infiltration. Sequencing revealed homology to *Leishmania mexicana* or *L. amazonensis*. Birthweight percentile was modestly lower in the PCR-positive group (predicted mean 53.8% vs. 56.5%, p = 0.76), while small for gestational age showed a non-significant trend toward higher prevalence among PCR-positive cases (prevalence ratio = 2.06, 95% CI: 0.32–13.39, p = 0.45).

**Conclusions/Significance:** Subclinical, dynamic transmission of *Leishmania* parasites typically associated with cutaneous disease was detected in this endemic zone. The presence of *L. mexicana* in human placentas was confirmed by immunofluorescence and sequencing, without an associated inflammatory response. These findings highlight the potential of CL-associated *Leishmania* species to reach the placenta and buccal mucosa of newborns, warranting further epidemiological investigation into the consequences of vertical transmission in regions with endemic CL.

**Author summary:** Leishmaniasis is a parasitic infection mainly known for causing skin ulcers (cutaneous leishmaniasis) or severe internal disease (visceral leishmaniasis). While congenital transmission of *Leishmania* has been documented in visceral disease, the potential for cutaneous species to cross the placenta has received less attention. We studied 53 pregnant women living in a highly endemic region of Tabasco, Mexico, and found that one-third harbored *Leishmania* DNA in their placentas, while about one-fifth of their newborns showed positive buccal swabs. Importantly, the women showed no clinical signs of cutaneous leishmaniasis, indicating that subclinical infection can still allow parasites to reach the placenta. We confirmed the presence of the parasite in placental tissue using immunofluorescence microscopy and DNA sequencing, which identified the species as *Leishmania mexicana*. The parasite was found inside immune cells (macrophages), without causing visible inflammation, suggesting the parasite can persist silently in placental tissue. These findings are important for women of reproductive age living in or travelling to cutaneous leishmaniasis-endemic regions, as they and their newborns may be at risk of subclinical exposure to the parasite.

## Introduction

Leishmaniasis is a protozoal infection caused by *Leishmania* parasites and is traditionally classified into visceral leishmaniasis (VL) and cutaneous leishmaniasis (CL). VL is characterized by hepatosplenomegaly, fever, anemia, and weight loss, whereas CL manifests as granulomatous or ulcerative skin lesions. Globally, an estimated 900,000 to 1.6 million new cases occur each year [1]. Transmission occurs through the bite of infected sandflies, primarily *Lutzomyia* in the Americas and *Phlebotomus* in the Old World. In endemic regions, up to one in 50 sandflies may harbor *Leishmania* [2]. Laboratory studies suggest female sandflies have short adult lifespans, typically fewer than 20 days, though natural populations may live considerably longer [3]. Mature females require a blood meal during each oviposition cycle; with inter-bite intervals averaging approximately 6 days, they typically acquire parasites from only a small number of blood meals over their lifespan [3]. Although cutaneous lesions are recognized in humans and animals, they appear relatively infrequently, raising the question of potential subclinical infection.

The fact that *Leishmania* parasites have been detected in the blood of both symptomatic patients and asymptomatic individuals living in endemic areas of CL [4] suggests that CL infection is not limited to skin alone. Animal models provide additional evidence supporting this observation: *Leishmania* parasites may disseminate to distant tissues [5], including the placenta, as demonstrated in a murine model [6]. This finding challenges the traditional dichotomy in which VL is viewed as strictly systemic and CL as strictly cutaneous. The placenta presents a unique opportunity to investigate subclinical infection due to its high degree of vascularization and routine expulsion after delivery. It receives approximately 0.5 litres of blood per minute, meaning that the total blood volume of an adult circulates through it every ten minutes [7]. This extensive circulation enables continuous exchange of nutrients, immune cells, and potentially pathogens between the mother and fetus. During passage through the placenta, some parasites may cross the barrier, a phenomenon well described for malaria, Chagas’ disease, and toxoplasmosis, which has been linked to adverse maternal and infant outcomes including anemia, low birth weight, pre-eclampsia, restricted fetal growth, and infant mortality [8, 9].

Given these parallels, it is plausible that *Leishmania* may also cross the placenta, leading to adverse pregnancy outcomes [10]. Evidence supporting vertical transmission of *Leishmania* has been described in VL, where both symptomatic and asymptomatic mothers have been shown to transmit the parasite to their offspring [11, 12, 13]. However, studies on the vertical transmission of CL-associated species remain scarce, although their findings are suggestive. In a Brazilian cohort of pregnant women with CL (n = 26), 10.5% experienced preterm birth and 10.5% had stillbirths, both associated with exuberant lesions [14]. Additionally, two toddlers from south-eastern Mexico were diagnosed with congenital visceral leishmaniasis caused by *L. mexicana* despite their mothers having no prior history of cutaneous lesions [15]. Nevertheless, the prevalence of *Leishmania* infection (clinical or subclinical) among women of reproductive age, and the effects of locally endemic CL-associated strains such as *L. mexicana* on birth outcomes, remain largely unknown. Given that suboptimal fetal growth is six times more prevalent in developing regions [16], many of which overlap with leishmaniasis-endemic zones, it is important to determine whether any relationship exists between them.

Protozoan infections (malaria, toxoplasmosis, Chagas’ disease, and visceral leishmaniasis) are well-documented causes of adverse fetal development, prompting public health efforts to enhance screening, diagnosis, and treatment [17, 18]. In contrast, leishmaniasis, particularly CL-associated strains, has not received the same level of attention as a potential threat to perinatal health. No study has systematically investigated the presence of *Leishmania* in placental tissue from women in CL-endemic regions, alongside the buccal mucosa of their newborns, or its potential association with adverse perinatal outcomes.

We conducted an exploratory cross-sectional study in a highly endemic zone of Tabasco, Mexico, with three specific aims: (1) to determine the frequency of placental infection by *Leishmania* sp. by PCR analysis, hypothesizing that subclinical infection by CL-associated species could be identified in the placentas of women living in endemic zones; (2) to ascertain the frequency of *Leishmania* sp. infection in the oral mucosa of newborns, hypothesizing that vertical transmission could occur either hematogenously or via parasite-harboring amniotic fluid reaching the buccal mucosa of the fetus; and (3) to explore the association of placental *Leishmania* infection with fetal growth, hypothesizing that infection would be associated with fetal growth restriction.

## Materials and Methods

### Study Design and Setting

We conducted a cross-sectional exploratory study in Tabasco, Mexico, a State in the south-east of the country encompassing some of the highest CL-endemic zones in Mexico. Highly endemic zones, defined as areas with more than 200 cases per 10,000 inhabitants in the preceding five years, were selected for sampling. Study zones were selected on the basis of the availability of National Health Services (*Secretaría de Salud*) facilities and accessibility by road (Figure 1).

**Figure 1.**
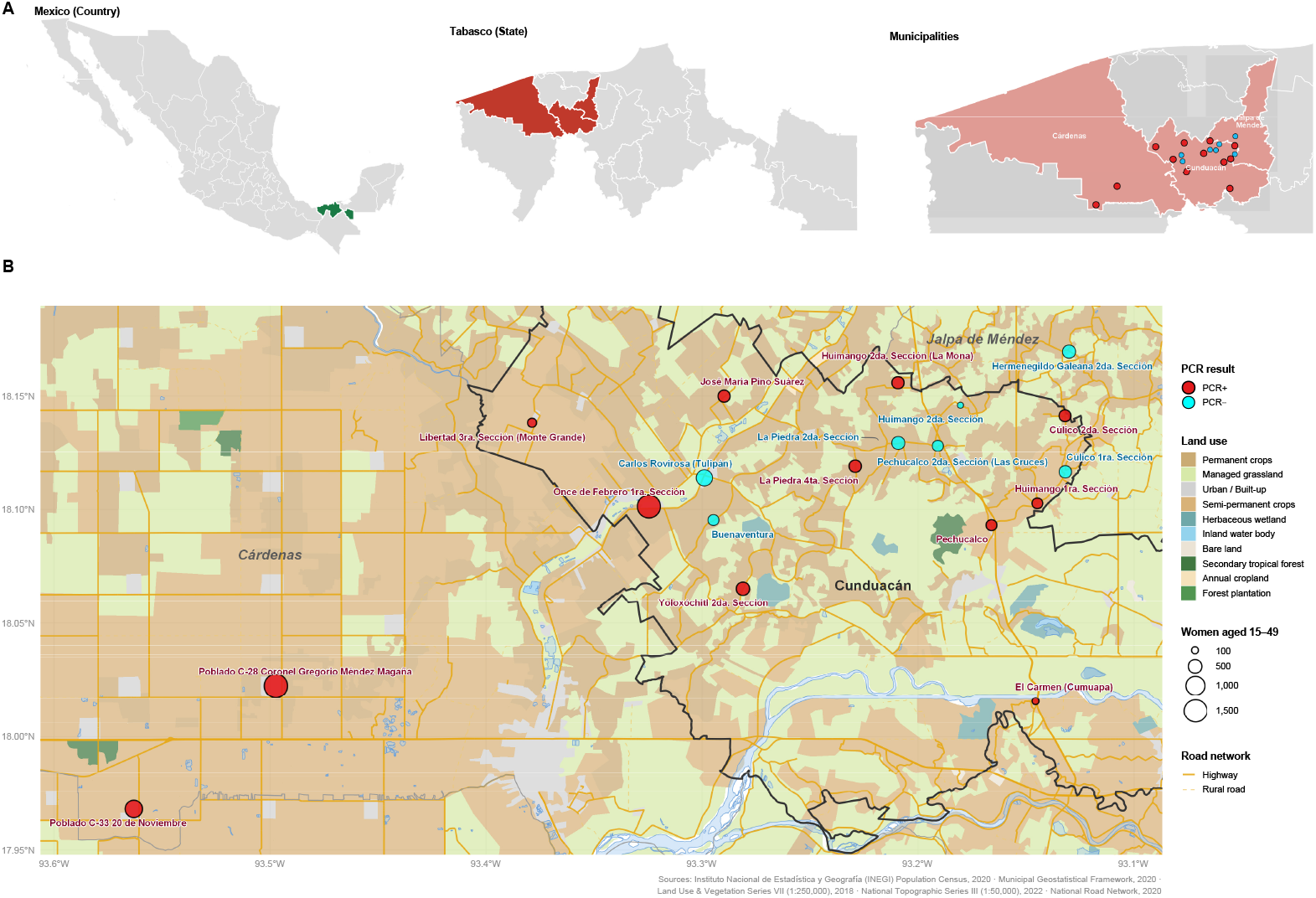
Regional distribution of placental leishmaniasis (PCR-positive) cases and land use in Cunduacán and neighboring municipalities (Tabasco, Mexico, 2017–2019). (A) Geographic location of the study area within Mexico. (B) Spatial distribution of sampled localities by placental PCR status. Each circle represents a sampled locality centroid from the INEGI 2020 Geostatistical Framework, the smallest geographic unit reported in publicly available census data; circle size is proportional to the number of women aged 15–49 years per locality (INEGI Census 2020). Red circles indicate localities with at least one PCR-positive placental sample; cyan circles indicate localities where all samples were PCR-negative.

Three secondary-level regional hospitals located in Cárdenas, Comalcalco, and Paraíso, and two tertiary-level hospitals (*Hospital de la Mujer* and *Hospital Rovirosa*) where women with complicated pregnancies are referred, were invited to participate. Community health houses (*casas de salud*) managed by the Tabasco Health Services were used as recruitment centers for pregnant women beginning at their first prenatal visit. Participating women then delivered at the corresponding hospital facility.

### Participant Recruitment

Women were eligible for inclusion if they: resided in a highly endemic CL zone, were in their first trimester of pregnancy, carried a singleton fetus, and received prenatal care at one of the participating *Secretaría de Salud* facilities. Women were excluded if they had multiple fetuses, a molar pregnancy, a history of toxoplasmosis, malaria, rubella, HIV, or exposure to other known teratogens, or a fetal trisomy. A total of 53 pregnant women meeting the inclusion criteria were recruited between April 2018 and April 2020.

The study protocol was approved by the Institutional Review Boards of the Instituto Nacional de Perinatología (#2017-3-116), the *Secretaría de Salud de Tabasco* (#INV/2133/PCI/1116), and Indiana University (#1905780741). All participants provided written informed consent during prenatal consultations. As the legal guardians of the neonates, mothers granted written consent for their newborn children’s participation, covering placental sampling, buccal swab, and urine collection performed at delivery. A red dot label was placed on clinical records after providing consent to identify study participants as placental donors at hospital arrival for delivery. All participants received standard-of-care prenatal and delivery care at no cost, as provided within the public health system, at a secondary or tertiary care hospital in the study area. PCR-positive results were not treated as clinical cases of leishmaniasis. The detection of parasite DNA is not equivalent to active clinical infection; furthermore, the drugs used to treat clinical leishmaniasis carry significant risks including hepatotoxicity and nephrotoxicity that outweigh likely benefits in the absence of clinical disease. Quarterly technical reports were submitted to health authorities in Tabasco to ensure follow-up monitoring of mothers and infants for potential CL lesions or clinical signs of vertical transmission.

### Data Collection

Nurses, doctors, and students involved in the care of pregnant women in the endemic area received training on project objectives, sample collection procedures, and participant communication. Quarterly refresher sessions were conducted to address questions and reinforce the training.

#### Medical Records

Data on maternal age, reproductive history (parity, prior miscarriages, prior stillbirths, prior Caesarean section), medical history (obesity, smoking, alcohol use, and drug use), prenatal clinical and laboratory findings (hypertension, gestational diabetes, hemoglobin), and previous or current *Leishmania* skin lesions were obtained by chart review of hospital records by researchers. Anemia in pregnancy was defined as a hemoglobin concentration in blood lower than 11.0 g/dL [19].

#### Perinatal Outcomes

Newborn data obtained from medical records included estimated gestational age in weeks (as determined by the best available dating method, with ultrasound-based dating and delivery-time registration in agreement), anthropometric measurements including birthweight (g) and crown-heel length (cm). Birthweight percentile for gestational age and sex was calculated using the INTERGROWTH-21st standard [20]. Small-for-gestational age (SGA) was defined as birthweight below the 10th percentile for gestational age and sex.

#### Placental Samples

At delivery, trained medical personnel collected four placental tissue samples of approximately 1 cm3 each from every participant for histological analysis and PCR testing. Placentas were sampled from four adjacent locations, two at the border and two close to the umbilical cord (Figure 2). Samples intended for histological analysis were stored in 1% formaldehyde (Sigma-Aldrich, USA). Samples intended for DNA extraction, PCR, or sequencing were stored in RNA Protect Tissue Reagent (Qiagen, Hilden, Germany).

**Figure 2.**
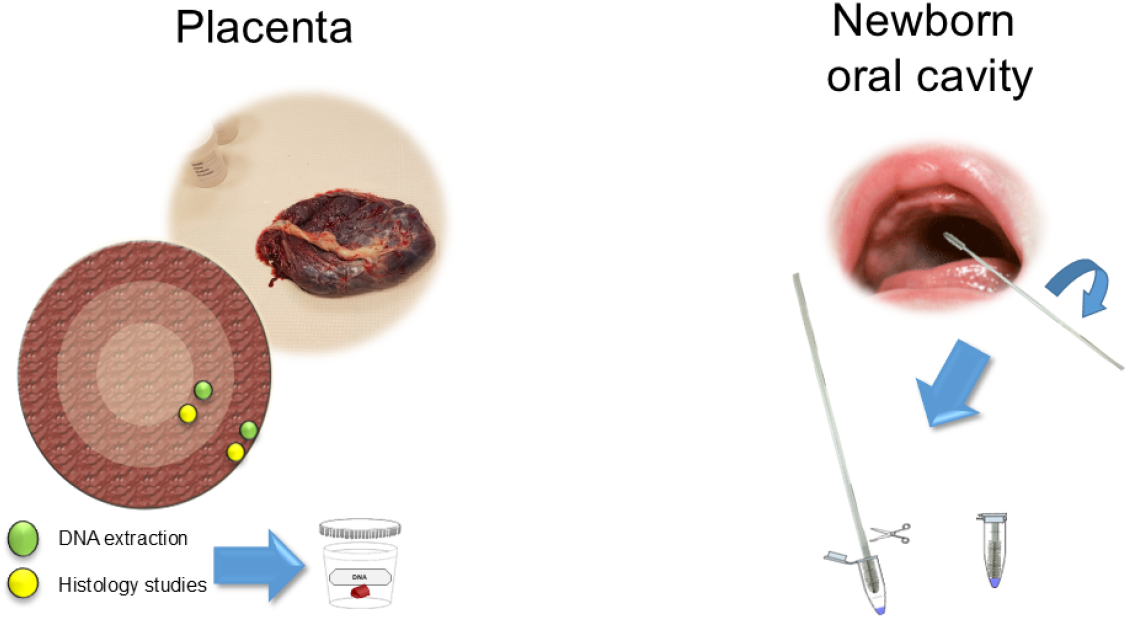
Sampling processes. Placentas were sampled from four adjacent locations, two at the border and two close to the umbilical cord. The dots in green represent those used for DNA extraction and the PCR procedure. Dots in yellow were fixed for histology studies. Newborn oral cavity samples were collected soon after birth.

#### Buccal Samples

Trained medical personnel collected a buccal swab from each newborn within the first few minutes after delivery and before their first feeding (Figure 2). Buccal swabs were preserved in 300 µL of Cell Lysis Solution (Gentra Puregene Buccal Cell Kit, Qiagen, Hilden, Germany). All laboratory testing was performed at the Instituto Nacional de Perinatología in Mexico City.

### Laboratory Analysis

#### DNA Extraction

Placental tissue was disrupted using a Tissue Lyser LT (Qiagen, Hilden, Germany) with steel beads (Qiagen, Hilden, Germany) for 20 min at 40 Hz. DNA extraction was then performed using a QIAamp DNA Mini Blood Kit (Qiagen, Hilden, Germany) following the manufacturer’s protocol. Buccal swabs were processed using a Gentra PureGene Buccal Cell Kit (Qiagen, Hilden, Germany) per the manufacturer’s recommendations.

#### PCR Amplification

DNA amplification for *Leishmania* sp. detection used the kinetoplast minicircle primers JW11 (forward, 5’-CCTATTTTACACCAACCCCCAGT-3’) and JW12 (reverse, 5’-GGGTAGGGGCGTTCTGCGAAA-3’). These primers amplify an approximately 120-bp fragment of the minicircle kDNA of *L. major*, which contains ∼10,000 copies per parasite [21]. End-point PCR used Taq PCR Master Mix (Qiagen, Germany) in a final volume of 25 µL. Cycling conditions were: initial denaturation at 95°C for 10 min; 40 cycles of denaturation at 95°C for 1 min, annealing at 58°C for 30 sec, and elongation at 72°C for 30 sec; and final extension at 72°C for 10 min (Figure 3).

**Figure 3.**
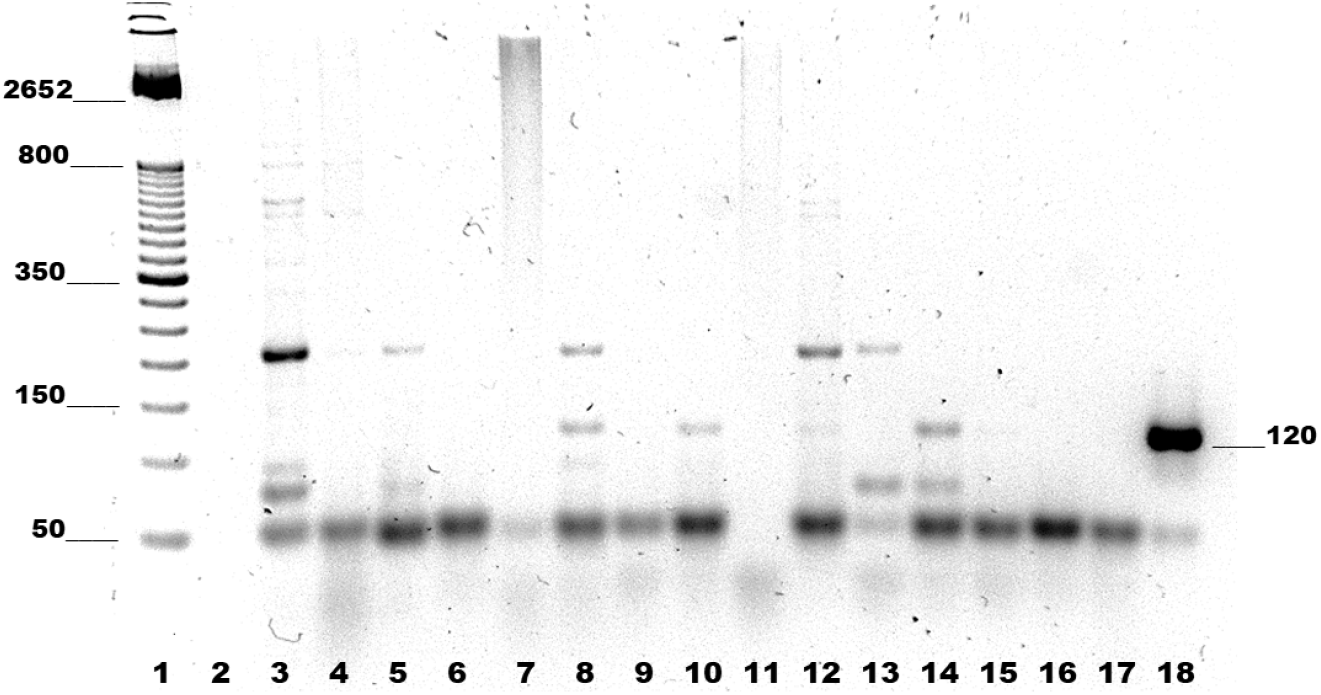
Agarose gel electrophoresis of placental samples subjected to endpoint PCR amplification. Lane 1: molecular weight marker; lane 2: reagents (no-template) control; lanes 3–16: paired border and inner placental samples; lane 17: *Trypanosoma cruzi* DNA as a negative amplification control; lane 18: *Leishmania mexicana* DNA as positive control, showing the expected 120-bp band amplified with JW11–JW12 primers. Lanes 3–4: term labor sample from a non-endemic zone, without the 120-bp band. Lanes 5–8 and 13–16: term pregnancy samples from endemic zone; the 120-bp band is present in lanes 8 and 14 (placenta border). Lanes 9–12: term pregnancy samples from two women with high-risk pregnancies who delivered at a tertiary-level hospital; the 120-bp band is visible in lanes 10 and 12.

#### Controls

Positive controls included *Leishmania mexicana* (MNYC/BZ/62/M379) and *Leishmania mexicana* (MHOM/MX/01/Tab3), as well as genomic DNA from *Trypanosoma cruzi* (*ninoa mexicana*), *Toxoplasma gondii* strain RH ATCC^®^ 50174D, and *Plasmodium falciparum* Welch Strain 3D7 Genomic DNA ATCC^®^ PRA-405D^™^. Negative controls consisted of reactions without a DNA template. The integrity of the extracted DNA was verified by 1% agarose gel electrophoresis and confirmed through amplification of the human *β*-globin gene.

#### Sequencing

Two sequencing protocols were followed to confirm the identity of amplified DNA. PCR-amplified bands were visualised by agarose gel electrophoresis, extracted with a QIAquick Gel Extraction Kit (Qiagen, Germany), and sent to the Instituto de Biotecnología of the Universidad Nacional Autónoma de México (UNAM) for sequencing. Secondary sequencing was also performed for the internal transcribed spacer 1 (ITS1) and 5.8S ribosomal gene regions, a validated molecular target for *Leishmania* species identification [22, 23], at the Indiana University-Bloomington Global Environmental Health Laboratory. Bioinformatic analyses compared sequences with GenBank database records via the Basic Local Alignment Search Tool (BLAST) at the National Center for Biotechnology Information (NCBI; http://www.ncbi.nlm.nih.gov).

#### Histology

For immunofluorescence detection of *Leishmania* in placental tissues, tissue sections were fixed on glass slides. Antibodies were purified from a patient with diffuse cutaneous leishmaniasis from Comalcalco, Tabasco, using a Protein G column (Sigma-Aldrich, St. Louis, MI, USA). Antibodies were labeled using the Mix-n-Stain CF 488A Antibody Labeling Kit (Sigma-Aldrich) per the manufacturer’s instructions and used at a 1:50 dilution. Counterstaining was performed using a 5 µg/mL solution of propidium iodide (Thermo Fisher Scientific, Baltics, UAB) for 3 min.

## Data Analysis

Descriptive statistics for maternal sociodemographic, reproductive, prenatal, and medical history characteristics, and infant outcomes are presented as n (%) for categorical variables, mean ± SD for normally distributed continuous variables, and median (IQR) for non-normally distributed continuous variables. Given the small and unequal group sizes, between-group comparisons used Welch’s t-test or Wilcoxon rank-sum test for continuous variables, and chi-square or Fisher’s exact test for categorical variables, as appropriate. Statistical significance was defined as p < 0.05.

The prevalence ratio (PR) of small for gestational age per placental PCR positivity was estimated using a modified Poisson regression model (log link) and robust standard errors with 95% confidence intervals (95% CI) [24]. To examine the association between placental PCR status and birthweight percentile, a fractional logit regression model (quasibinomial family with logit link) was fitted and presented as odds ratios (OR) with 95% CI. Predicted mean birthweight percentiles with 95% CI were estimated from the model and displayed graphically alongside observed data. Due to the small sample size, multivariable adjustment was not performed; all regression estimates are therefore unadjusted (crude).

The enrollment of 53 participants are the total number of women who met eligibility criteria and provided consent over the study period (April 2018–April 2020); no formal *a priori* power calculation was performed given the exploratory nature of the study.

For variables with missing data, complete-case analysis was used. Hemoglobin values were available for 34 of 53 participants (64%); reasons for missingness were not recorded. Analysis of buccal PCR status was based on 52 newborns, as one swab was not received. No additional sensitivity analyses were pre-planned given the exploratory study design.

All analyses were conducted in R (version 4.5.3) through R Studio (version 2026.01.1). The source code for this manuscript, statistical analyses, and project documentation is available through the online repository: https://github.com/javimangal/leishmania-placental.

## Results

### Study Population and PCR Analysis

Most (37 of 53; 96%) of the study participants delivered at the Regional Hospital of Cunduacán; the remainder gave birth at Villahermosa hospital sites (n = 2). Placental PCR was positive in 18 (34%) participants and negative in 35. The geographic distribution of PCR-positive cases across sampled localities is shown in Figure 1; positive cases were distributed across 13 out of 19 (68.4%) sampled localities.

The amplification of a 120-bp fragment, which appears as a unique product when DNA is amplified from *L. mexicana* reference strains as well as from regional isolates, was used as the criterion for a positive result. Both the internal (near the umbilical cord) and external placental regions underwent successful PCR analysis for all 53 maternal participants. The sampling scheme is illustrated in Figure 2. In five patients, both the internal and external placental regions were positive; the internal region tested positive in 13 samples (8 showing intense bands, 5 faint bands), while the external region tested positive in 10 samples (5 intense, 5 faint; Figure 3).

Of the 53 newborn buccal swabs successfully tested for DNA integrity, 11 (20.8%) tested positive by endpoint PCR. However, only 3 cases showed matching positive results for both the buccal swab and placental samples; 8 newborns with PCR-positive buccal swabs were born from mothers with PCR-negative placentas.

### Confirmation of Parasite Presence by Immunofluorescence

Histological analysis was performed on eight placentas (three PCR-positive and five PCR-negative). In all participants, histological examination revealed features consistent with term placentas. In PCR-positive placentas, macrophages containing fluorescent structures resembling intact amastigotes were observed. These parasitized fluorescent macrophages (pfMØs) were detected on both the maternal and fetal sides, ranging from the syncytiotrophoblast to the chorionic villi. The pfMØs were specifically located at the margins of fetal sinusoids and on the endothelium of the basal decidua (Figure 4). Fluorescent macrophages were also identified within the characteristic reticular stroma of the basal plate and intermediate villi, extending from the syncytiotrophoblast. In all cases, fluorescence was observed inside the macrophages without surrounding inflammatory cell infiltration, suggesting the absence of an active inflammatory response. No pfMØs were detected in PCR-negative placentas.

**Figure 4.**
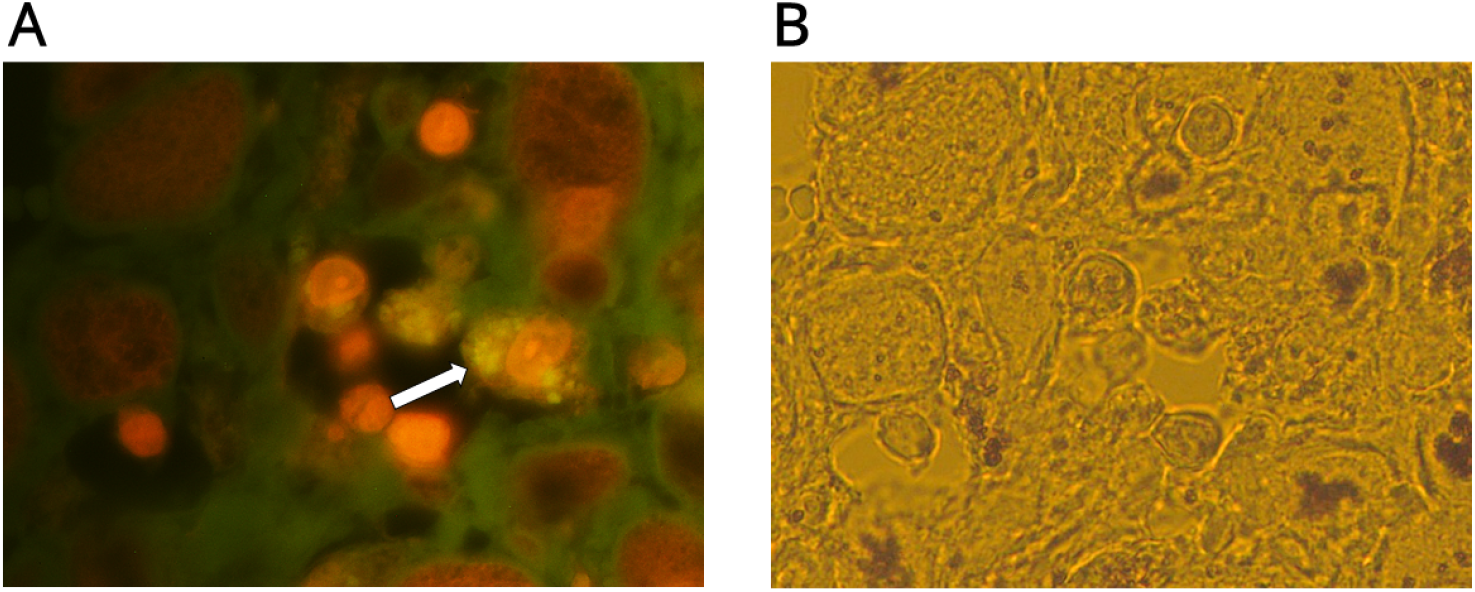
Histology of a PCR-positive placenta. Pictures of the same basal decidua field. (A) Brightfield image. (B) Staining with a primary FITC-conjugated anti-*Leishmania* antibody from a patient with diffuse cutaneous leishmaniasis and counterstaining with propidium iodide (PI). The arrow indicates a macrophage with fluorescent intracellular amastigotes. 1000× magnification.

### Confirmation and Identification of *Leishmania* by Sequencing

Three placentas with intense bands on the PCR agarose gel were selected for band elution and sequencing. The 120-bp forward product of the first sample showed 90% homology to *L. mexicana* (Z11556.1); this participant was a woman aged 30–34 at 39 weeks of gestation with hypertension who delivered a newborn in the 90^th^ weight percentile. The second sample showed 87% homology to *L. amazonensis* (EU370874); this participant was a woman aged 20–24 in her fourth pregnancy with a prior miscarriage who delivered at 40 weeks with a newborn also in the 90^th^ weight percentile. A third sample showed homology to *L. panamensis* (M87314.1); this participant was a woman aged 25–29 with a history of miscarriage and gestational diabetes who delivered a newborn in the 95^th^ weight percentile. All three samples presented sequence homology to a region within the minicircle of kinetoplast DNA. Separate amplification using a *L. mexicana* primer targeting the ITS1 and 5.8S ribosomal gene regions confirmed the presence of *L. mexicana* in placental samples (GenBank accession KU177477.1; 617 bp; 93–99% homology to reference strain M379).

### Maternal and Infant Characteristics by *Leishmania* PCR Status

Table 1 presents the comparison of maternal and infant characteristics by placental PCR status. Most participants (69.8%) were between 20 and 35 years of age. Mean (SD) maternal age was 24.6 ± 6.6 years overall and did not differ significantly between PCR groups (p = 0.16). Most participants had delivered at least once before (11.3% had ≥ 3 prior deliveries). Average parity did not differ between groups (p = 0.27). None of the participants presented with CL lesions. None reported tobacco, alcohol, or illicit drug use during pregnancy.

**Table 1.**
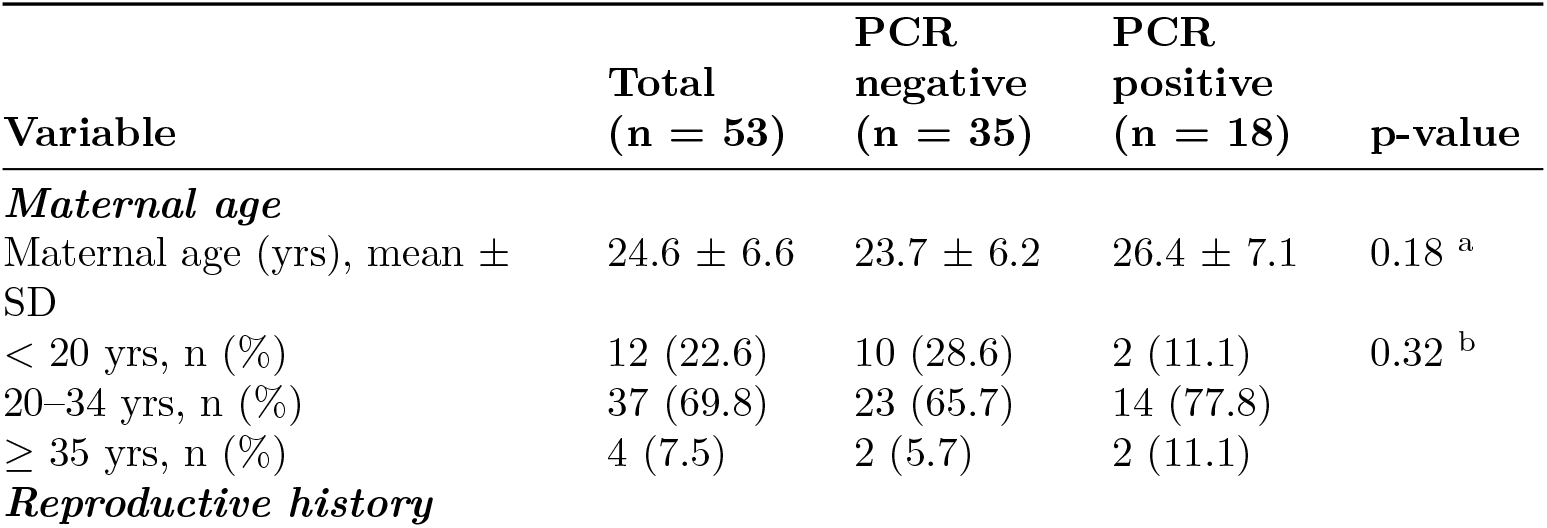
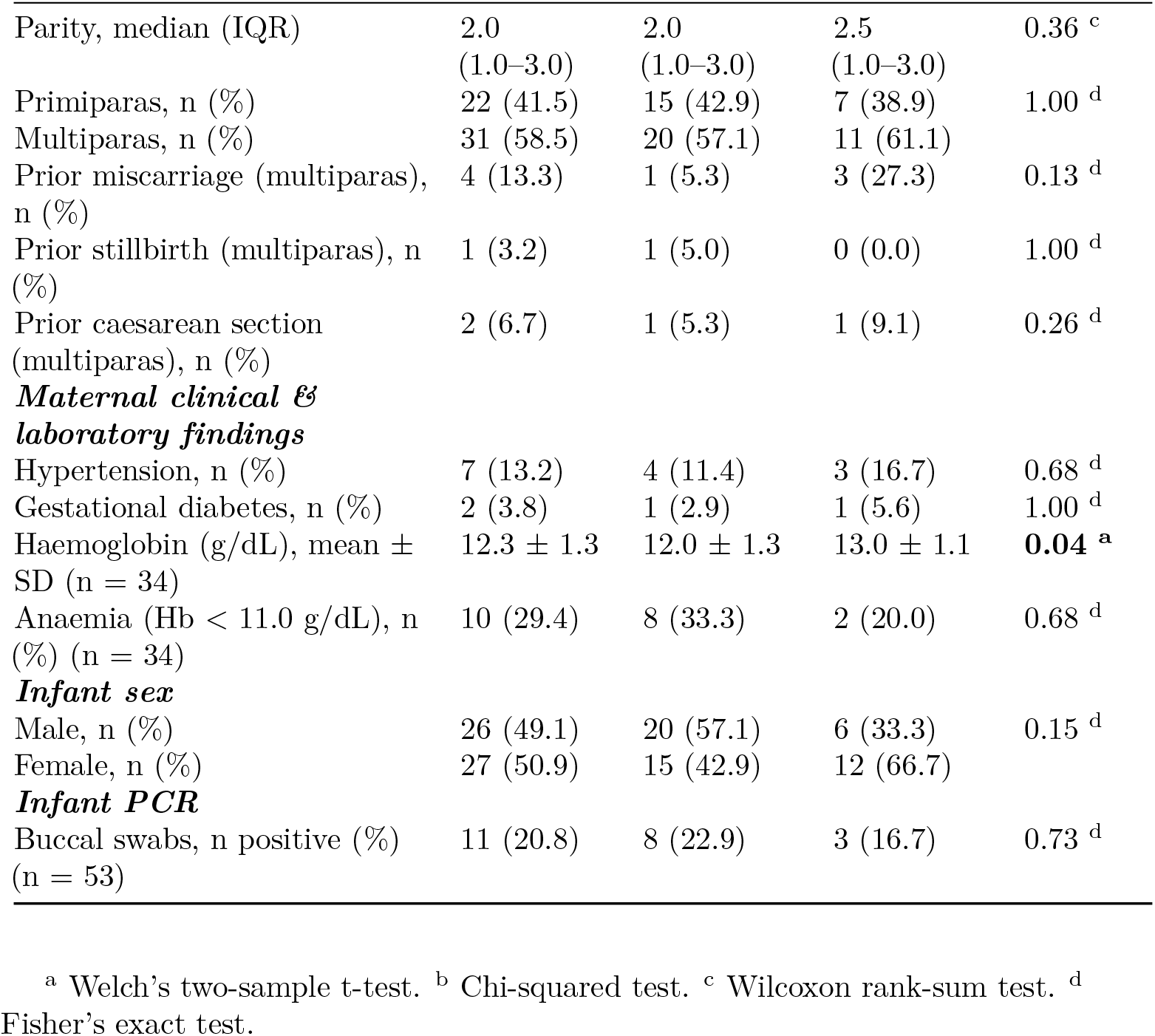
Comparison of maternal and infant characteristics by *Leishmania* PCR result in placental samples.

Among multiparas, prior miscarriage was more common in the PCR-positive group than in the PCR-negative group (3/11 [27.3%] vs. 1/20 [5.0%]; p = 0.13), although this difference was not statistically significant. The groups did not differ significantly with respect to prior stillbirth or prior Caesarean section. The proportion of male newborns was lower in the PCR-positive group (6/18, 33.3%) than in the PCR-negative group (20/35, 57.1%), though this difference was not statistically significant (p = 0.18).

### Perinatal Outcomes

Table 2 presents the comparison of perinatal outcomes by PCR status. Gestational age and birthweight did not differ significantly between groups (p = 0.19 and p = 0.60, respectively). Crown-heel length was slightly lower in the PCR-positive group (median 50.5 vs. 52.0 cm), though this difference was not significant (p = 0.28). Five-minute Apgar scores were uniformly 9 in both groups, with no infant scoring below 7. SGA was observed in 2/18 (11.8%) of the PCR-positive group versus 2/35 (5.7%) of the PCR-negative group (p = 0.59). The crude prevalence ratio for small for gestational age of PCR-positive cases compared to PCR-negative was PR = 2.06 (95% CI: 0.32–13.39; p = 0.450).

**Table 2.** Comparison of perinatal outcomes by *Leishmania* PCR result in placental samples.

| Variable | Total<br>(n = 53) | PCR<br>negative<br>(n = 35) | PCR<br>positive<br>(n = 18) | p-value |
| --- | --- | --- | --- | --- |
| **Perinatal outcomes** |  |  |  |  |
| Gestational age (wks),<br>median (IQR) | 39.0<br>(38.0–40.0) | 39.0<br>(39.0–40.0) | 39.0<br>(38.0–40.0) | 0.19 <sup>a</sup> |

| Variable | Total<br>(n = 53) | PCR<br>negative<br>(n = 35) | PCR<br>positive<br>(n = 18) | p-value |
| --- | --- | --- | --- | --- |
| Birthweight (g), median<br>(IQR) | 3320.0<br>(3000.0–<br>3500.0) | 3400.0<br>(3100.0–<br>3500.0) | 3220.0<br>(2862.5–<br>3552.5) | 0.60 <sup>a</sup> |
| Crown-heel length (cm),<br>median (IQR) | 51.0<br>(49.5–53.0) | 52.0<br>(50.0–53.0) | 50.5<br>(49.0–52.0) | 0.28 <sup>a</sup> |
| ***Birthweight***<br>***classification*** |  |  |  |  |
| Weight percentile, median<br>(IQR) | 55.0<br>(40.0–80.0) | 60.0<br>(45.0–80.0) | 50.0<br>(40.0–70.0) | 0.87 <sup>a</sup> |
| Small-for-gestational age (<<br>10th percentile), n (%) | 4 (7.7) | 2 (5.7) | 2 (11.8) | 0.59 <sup>b</sup> |
| ***Apgar score*** |  |  |  |  |
| 5-min. Apgar score, median<br>(IQR) | 9.0 (9.0–9.0) | 9.0 (9.0–9.0) | 9.0 (9.0–9.0) | 0.61 <sup>a</sup> |
| 5-min. Apgar score < 7, n<br>(%) | 0 (0.0) | 0 (0.0) | 0 (0.0) | - |

<sup>a</sup> Wilcoxon rank-sum test; <sup>b</sup> Fisher's exact test.
Abbreviations: MC = miscarriage; SB = stillbirth; SGA = small-for-gestational age.

### Birthweight Percentile by PCR Status

Placental PCR positivity was associated with a modest, non-significant reduction in birthweight percentile (Figure 5). The fractional logit model estimated lower odds of a higher birthweight percentile in PCR-positive compared to PCR-negative women (OR 0.9, 95% CI 0.45–1.78; p = 0.76). Model-predicted mean birthweight percentiles were 56.5% (95% CI 46.7–65.8) in the PCR-negative group and 53.8% (95% CI 39.9–67.1) in the PCR-positive group, corresponding to an absolute difference of 2.7 percentile points (95% CI −14.3–19.6).

**Figure 5.**
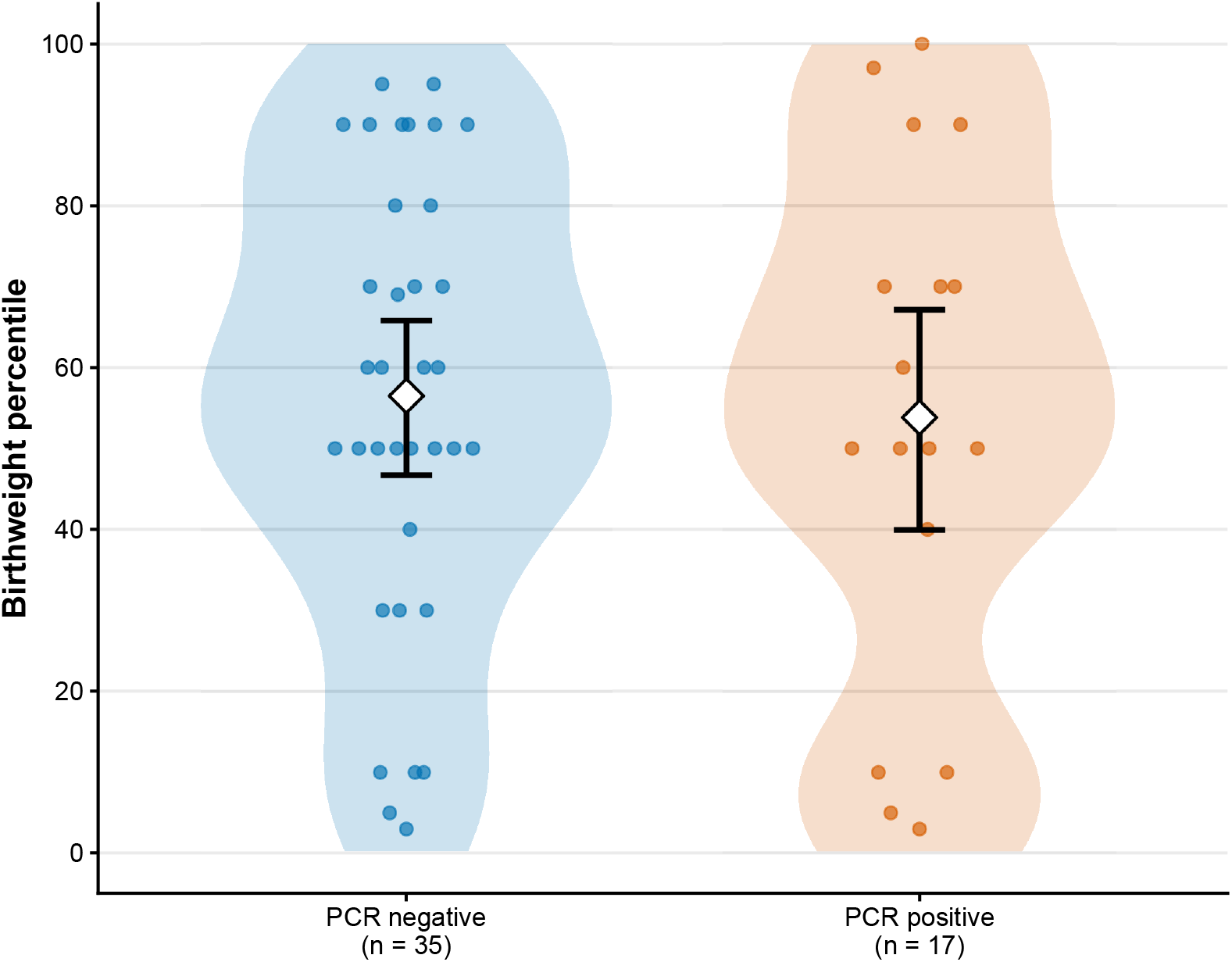
Birthweight percentile by placental PCR status. Points show individual observations; shaded areas correspond to the density distribution. White diamonds and bars show fractional logit predicted means with 95% confidence intervals.

## Discussion

The present findings confirm the presence of *Leishmania mexicana*, a species traditionally associated with cutaneous lesions, in a relatively high proportion of human placentas from CL-endemic regions of Mexico. One-third of placentas tested positive by PCR, and parasitized macrophages were confirmed by immunofluorescence in PCR-positive placentas, without an associated inflammatory response.

Congenital transmission of *Leishmania* is well documented for VL and is associated with adverse pregnancy outcomes, including fetal loss, low birth weight, and the subsequent development of visceral leishmaniasis in infants [25, 26]. For CL, such transmission has been poorly documented. Our results, together with the recent demonstration of vertical transmission of *L. donovani* in a mouse model of VL [11], add to the growing body of evidence that *Leishmania* parasites from both visceral and cutaneous species can reach the placenta.

For other hematological protozoans with congenital transmission such as *Toxoplasma, Trypanosoma*, and *Plasmodium*, adverse pregnancy effects have been predominantly associated with the first pregnancy, suggesting a protective effect of maternal immunity in subsequent pregnancies [27, 17]. In contrast, in our study perinatal complications were more common in multiparas (specifically prior miscarriage), although this difference was not statistically significant.

Compared with the high proportion of the population harboring *Leishmania* parasites in this endemic zone, the prevalence of clinical disease is relatively low. Several factors may explain this discrepancy, including the low pathogenicity of circulating *Leishmania* strains, low parasite load, or individual host susceptibility [27]. Additionally, circulating strains in this zone have been previously reported to undergo apoptotic destruction in vitro at physiological temperature (37°C) [28]; however, within the complex in vivo environment, the parasite’s ability to survive may differ, potentially because rescue molecules may influence its persistence.

The detection of parasites by PCR does not distinguish between live and dead parasites, and this study did not include parasite cultures from placental samples. A notable histological feature observed in this study, compared with infections caused by *T. cruzi* and *T. gondii*, was the absence of a host-stimulated immune response [29]. Unlike those protozoa, immune cells were not observed in the vicinity of mononuclear cells harboring *Leishmania* parasites, consistent with the parasite’s well-characterized ability to evade intracellular immune responses. In a mouse model of VL, IFN signal activation and cellular immune suppression have been shown to accompany placental *L. donovani* infection [11], further supporting the concept that *Leishmania* parasites may create an immunologically permissive environment within the placenta.

The detection of parasite DNA in one-fifth of newborn oral cavity samples suggests either contact with amniotic fluid harboring parasites or infection via the hematological route. In a mouse model infected with *L. mexicana* via the footpad, the parasite disseminated to all vascularized organs studied, including the salivary glands [5], supporting the plausibility of hematogenous transmission to the newborn’s oral mucosa. Notably, 8 of the 11 buccal PCR-positive newborns were born from PCR-negative placenta mothers, suggesting that the placenta is not the only potential route and that contamination at the time of delivery cannot be excluded. A longitudinal follow-up of this cohort could provide valuable insights into potential long-term effects on the offspring’s immune tolerance [13].

The minor sequence variations observed among the sequenced samples suggest the existence of genetically diverse circulating strains. Future studies should analyze the protozoal microbiome of the placenta and compare the genetic sequences of parasites detected in placental tissue with those from cutaneous lesions, to identify potentially non-pathogenic strains. For *T. cruzi* [30] and *Plasmodium* [31], specific parasite strains involved in congenital transmission have been identified; it is plausible that a similar pattern exists for *Leishmania*.

Reproductive-age women living in or travelling to tropical regions endemic for CL represent a high-risk population for adverse pregnancy outcomes and subclinical congenital transmission. In non-endemic areas, they may also serve as a potential source of *Leishmania* congenital transmission. The full consequences of such transmission remain unknown, as congenital CL has not been previously suspected. A key aspect to consider is the potential impact on central immune tolerance development in offspring exposed in utero, which warrants further investigation.

Limitations of this study include the cross-sectional exploratory design precluding causal inferences. The sample size was small, reducing statistical power to detect differences in perinatal outcomes; expansion of the original recruitment target was rendered infeasible by the COVID-19 pandemic. PCR positivity does not confirm the presence of viable parasites as PCR detects DNA from both live and dead organisms, which may overestimate the prevalence of active infection. The absence of parasite cultures is a further limitation in this respect. Hemoglobin data were available for only 34 of 53 participants (64%), limiting the assessment of anemia as an outcome; the reasons for missing data were not recorded. The generalizability of these findings to other CL-endemic regions should be approached with caution. Sandfly vector species, circulating *Leishmania* strains, and host immune responses may vary substantially across geographic areas, and these results may not be representative of all endemic populations.

## Conclusion

*Leishmania* DNA was detected in one-third of placentas from women living in CL-endemic zones with term pregnancies, confirming that dynamic subclinical transmission occurred below clinical expression levels. The presence of *Leishmania mexicana* in placental tissue was confirmed by direct immunofluorescence and sequencing. Notably, inflammatory infiltrates were not observed surrounding the mononuclear cells harboring the parasites, suggesting a clinically silent infection. Parasite DNA was also detected in the buccal mucosa of one-fifth of newborns. These findings call for increased awareness of the potential for CL-associated *Leishmania* species to undergo vertical transmission, with implications for maternal and neonatal health surveillance in endemic regions.

## Funding

This study was supported by funding from the National Perinatology Institute to Dr. Galindo-Sevilla (grant #2017-3-116) and from the Indiana University President’s International Research Award to Drs. Armijos and Weigel (grant #PSA-PRO-0680-2020; PIRA10302019). Dr. Berger was supported by the University of Chicago Pritzker School of Medicine Pritzker Fellowship and the American Society of Tropical Medicine and Hygiene Benjamin H. Kean Travel Fellowship. The institutional funders had no role in the study design, data collection, analysis, decision to publish, or preparation of the manuscript.

## Data Availability

The data supporting the conclusions of this article are available upon reasonable request to the corresponding author. The source code for the analyses and manuscript is publicly available in the GitHub repository: https://github.com/javimangal/leishmania-placental.

## Acknowledgments

We are indebted to the pregnant women for their participation in this study. We are grateful to the Dirección de Calidad y Enseñanza de la Secretaría de Salud de Tabasco, as well as to the communities and hospital centres of Cunduacán, Comalcalco, Cárdenas, and Villahermosa for their cooperation. We also thank personnel at Jurisdicción Sanitaria #6, Cunduacán, Tabasco, for their support in participant recruitment. We are particularly grateful to Dr. Ángel Ernesto Chamdomid Salud, Director of the Hospital Regional de Cunduacán, for his institutional support, and to nurses Mauricio Mosqueda Fernández, Adelaida Rodríguez Tiquet, and Saúl Pérez for their invaluable assistance with sample collection and storage. Liliana Aline Fernández-Urrutia, Frida Häberle, and Miriam Torres-Vasquez acknowledge the Programa Nacional de Servicio Social en Investigación en Salud, Dirección General de Calidad y Educación en Salud (DGCES), for enabling their participation in this research at their respective institutions.

## Declaration of interests

The authors declare no competing interests.

## Author Contributions

Conceptualization: Norma Galindo-Sevilla, Brandon A. Berger, M. Margaret Weigel.

Data curation: Norma Galindo-Sevilla, Liliana Aline Fernández-Urrutia. Formal analysis: Norma Galindo-Sevilla, Rodrigo X. Armijos, Liliana Aline Fernández-Urrutia, Javier Mancilla-Galindo, M. Margaret Weigel, Frida Häberle.

Funding acquisition: Norma Galindo-Sevilla, Rodrigo X. Armijos, M. Margaret Weigel.

Investigation: Norma Galindo-Sevilla, Alejandra González Ayala, José Luis Acosta-Patiño, Esmelin Trinidad-Vázquez, Elizabeth Baz-Rojas, Candelaria Frías Selván, Juanita Ortíz-Avalos, Eva Cristina Avalos-Ortiz, Miriam A. Delgado-Hernandez, Liliana Aline Fernández-Urrutia, Frida Häberle, Miriam Torres-Vasquez, Yuriria Paredes, Miroslava Avila-García, Magdalena Aguirre García.

Methodology: Norma Galindo-Sevilla, Alejandra González Ayala, M. Margaret Weigel.

Project administration: Norma Galindo-Sevilla, José Luis Acosta-Patiño.

Resources: Norma Galindo-Sevilla, Rodrigo X. Armijos.

Software: Javier Mancilla-Galindo, Liliana Aline Fernández-Urrutia.

Supervision: Norma Galindo-Sevilla, Rodrigo X. Armijos, M. Margaret Weigel, Allison H. Bartlett.

Validation: Norma Galindo-Sevilla.

Visualization: Javier Mancilla-Galindo, Liliana Aline Fernández-Urrutia, Frida Häberle, Miriam Torres-Vasquez.

Writing – original draft: Norma Galindo-Sevilla, Rodrigo X. Armijos, M. Margaret Weigel.

Writing – review & editing: All authors.

## Notes

### Competing Interest Statement

The authors have declared no competing interest.

### Funding Statement

Yes

### Author Declarations

The study protocol was approved by the Institutional Review Boards of the Instituto Nacional de Perinatología (#2017-3-116), the *Secretaría de Salud de Tabasco* (#INV/2133/PCI/1116), and Indiana University (#1905780741). All participants provided written informed consent during prenatal consultations. As the legal guardians of the neonates, mothers granted written consent for their newborn children's participation, covering placental sampling, buccal swab, and urine collection performed at delivery.

